# High Prevalence of *Plasmodium* Infections and Low *P. malariae* msp-1 Diversity in Beninese Population: Implications for Malaria Surveillance and Vaccine Development

**DOI:** 10.64898/2025.12.23.25342548

**Authors:** Romuald Agonhossou, Romaric Akoton, Simon Rothbauer, Macqueen Ngum Mbencho, Victorine M. Ndiwago, Thirumalaisamy P. Velavan, Ayola A. Adegnika

## Abstract

**Background:** *Plasmodium malariae,* one of the five parasite species causing malaria in humans, though less widespread than *P. falciparum*, can result in prolonged and persistent infections. This research aims to monitor asymptomatic *Plasmodium* infections and characterize genetic diversity in the merozoite surface protein (msp-1) of *P. malariae* in Beninese population.

**Methods and findings:** Blood samples collected from 484 asymptomatic participants in Southern Benin underwent analysis for all human *Plasmodium* species utilizing rapid diagnostic tests (RDT - PfHRP2/LDH), microscopy, nested PCR, and nested real-time qPCR assay. Mono- or mixed infections of *P. malariae* underwent *msp-1* genotyping. The *P. malariae msp-1* gene was amplified by a nested PCR, followed by sanger sequencing.

Microscopy and RDTs identified infection rates of *Plasmodium* spp. at 17% and 20%, respectively, whereas nested PCR significantly elevated rates to 38% (P<0.0001). Furthermore, the utilisation of nested qPCR revealed a 12% increase in *Plasmodium* spp. prevalence among individuals who tested negative using nested PCR. Molecular methods notably enhanced the detection of *P. malariae* compared to microscopy, with prevalence rates of 7% and 12% using nested PCR and qPCR, respectively (P<0.05). Genetic analysis of *msp-1* revealed a high nucleotide sequence similarity (95-100%). The mean pairwise nucleotide divergence (π) of *P. malariae* was 0.00602. Gender-based analysis indicated a marginal risk of *P. malariae* infection in females compared to males (RR: 1.3, 95% CI: 0.9 - 8; P = 0.04).

**Conclusions:** The present study uncovered a high prevalence of *Plasmodium* infections, including sub-microscopic *P. malariae*. Furthermore, our findings demonstrated a low sequence diversity of *P. malariae msp-1* compared to other *Plasmodium* species. This suggests that msp-1 holds promise as a vaccine candidate against *P. malariae* infection, given its low genetic diversity and highly conserved amino acids.

**Author Summary:** Malaria remains a major public health challenge in sub-Saharan Africa, with most research focused on *P. falciparum*. However, non-falciparum species such as *P. malariae* are often overlooked despite their ability to cause prolonged and persistent infections. In this study, we investigated the prevalence of Plasmodium infections among 484 asymptomatic individuals in Southern Benin, with a particular focus on *P. malariae*. Using a combination of rapid diagnostic tests, microscopy, nested PCR, and nested quantitative PCR, we found that molecular methods significantly improved the detection of *Plasmodium* infections, revealing that *P. malariae* infections were more common than previously detected by conventional methods. Genetic analysis of the *msp-1* gene of *P. malariae* showed low sequence diversity, indicating highly conserved regions that could serve as potential vaccine targets. Our findings highlight the underestimated prevalence of *P. malariae* and demonstrate the utility of sensitive molecular tools for detecting sub-microscopic infections. These results provide important insights into the epidemiology and genetic diversity of *P. malariae* in Benin and support further research into vaccine development and malaria control strategies targeting non-falciparum species

## Introduction

Malaria remains a significant global health challenge, particularly in endemic regions [1]. Despite efforts, 282 million cases and 610,000 deaths occur annually, with sub-Saharan Africa accounting for 94% of the disease burden [2]. The WHO aims for universal access to malaria prevention, treatment, and diagnosis [3], yet many infections go untreated due to reliance on symptomatic diagnosis [4]. Effective malaria control necessitates comprehensive community-based strategies that specifically address asymptomatic carriers across all malaria species.

Of the five human *Plasmodium* species implicated in malaria transmission (P*. falciparum, P. vivax, P. malariae, P. ovale spp., and P. knowlesi*) [5], *P. malariae* presents distinct challenges due to its unique characteristics, including a 72-hour erythrocytic cycle and the ability to maintain low parasitaemia levels over extended periods [6,7]. In areas where other *Plasmodium* species are dominant, traditional diagnostic techniques like microscopy and rapid diagnostic tests (RDTs) often fall short in detecting *P. malariae* [8]. However, molecular methods such as PCR and qPCR could provide more accurate prevalence rates for *P. malariae* [6,9]. Given escalating worries over rising *P. malariae*, recent research indicates that untreated *P. malariae* infections may result in heightened mortality rates and severe complications, including nephrotic syndrome and severe anaemia [10–12]. Persistent detection of *P. malariae* has been documented in endemic regions for several decades. Studies conducted in West Africa revealed a rise in *P. malariae* prevalence from 1% to 13% between 2007 and 2010 in Burkina Faso [13], and from 1% to 2% between 2010 and 2019 in Benin [14]. Similarly, research from East Africa, particularly Tanzania, indicated a twofold increase from 2010 to 2016 [15].

While malaria vaccines show promise for malaria elimination, research has predominantly focused on genes that target different stages of the parasite’s life cycle [16,17]. Among these, merozoite surface protein 1 (msp-1), present on the surface of the merozoite (the parasite’s invasive form), has received considerable attention [17]. Extensive studies have explored its role in eliciting antibodies capable of impeding the parasite’s invasion of red blood cells, thereby preventing infection progression, and reducing malaria severity [18,19]. Additionally, *msp-1* demonstrates genetic variability[20,21], with diverse allelic forms (within parasite), making it appropriate for investigating strain-specific immune responses [22]. Moreover, its effectiveness against other malaria species such as *P. malariae* remains relatively unexplored, underscoring the necessity for additional research on *P. malariae* msp-1 diversity in malaria endemic areas.

This study aims to investigate *Plasmodium* infections in Beninese population using conventional microscopy and rapid diagnostic tests (RDT - PfHRP2/LDH). In addition, nested PCR and qPCR were used to improve the detection of all sub-microscopic *Plasmodium* species. The samples that tested positive for *P. malariae* were further characterized for *P. malariae* msp-1 genetic diversity.

## Materials and Methods

### Ethics approval

Ethical approval for this study was obtained from the Ethics Committee of the Faculty of Sciences and Health and the health authorities of the Ouidah-Kpomasse-Tori Bossito district (N°115/2018/CER-ISBA/FSS/UAC of 29 October 2018). A written informed consent form was obtained from the participating residents. The consent form and survey were signed/completed by a parent for participants under the age of 18. Malaria-positive residents were treated with artemisinin combination therapies (ACTs) according to the National Malaria Control Programme (NCMP) guidelines.

### Study population

This cross-sectional investigation took place within the rural regions of Ouidah and Kpomasse municipalities, situated in the southern region of Benin, during the dry seasons of January to March 2022 and March 2023. Detailed descriptions of the study areas have been previously documented [23,24]. Selection of the rural sites was based on preliminary findings regarding *P. malariae* prevalence in the study region [14]. A structured questionnaire was administered to collect socio-demographic and anthropometric data, including age, gender, weight, and height. Additionally, approximately 2 mL of venous blood was collected from each participant in an EDTA tube. This blood was utilized for various measurements, including haemoglobin levels, rapid diagnostic tests (RDTs) for *Plasmodium* spp. detection, preparation of thick and thin blood smears for microscopy, and preparation of filter paper samples for further molecular analysis.

### Diagnostic and genotyping procedures

#### Rapid diagnostic test

Rapid Diagnostic Test (RDT): The Bioline® Malaria 4 species test was utilized for rapid malaria detection, differentiating between *P. falciparum*, *P. malariae*, *P. vivax*, and *P. ovale*. The test differentiates between *P. falciparum* and *P. malariae*, *P. vivax*, and *P. ovale* based on the presence of specific proteins. The procedure involved using 5μL of whole blood from the EDTA tube and transferring it to the sample well. Following the manufacturer’s protocol, assay diluent was added, and results were read after 15 minutes.

#### Plasmodium spp. detection using Microscopy

Thin and thick blood smears were prepared, stained with Giemsa, and examined under a microscope for parasite identification and density determination following WHO recommendations [25].

#### Nested PCR for Plasmodium species identification

DNA was extracted from filter paper samples using commercial kits (Qiagen, Hilden, Germany) following the manufacturer’s instructions. The extracted DNA underwent nested PCR targeting the 18S rRNA gene region of Plasmodia, as described previously [26]. Species-specific primers were used in nested PCR to identify *P. falciparum*, *P. malariae*, *P. ovale*, and *P. vivax* as described previously. The PCR products were visualized by gel electrophoresis on a 1.5 % agarose gel stained with SYBR Green. Details of the primers used, and their expected size are provided in **Supplementary Table 1.**

#### Nested qPCR for Plasmodium species identification

Samples negative for nested PCR underwent nested qPCR in a two-step reaction as described previously [27] on a LightCycler480-II (Roche, Mannheim, Germany). The first step involved conventional PCR amplification using conserved primers for all plasmodia species [26] followed by qPCR using species-specific primers and probes for five human malaria species including *P. falciparum*, *P. malariae*, *P. vivax, P. ovale curtisi* and *P. ovale wallikeri*. Positive results were determined based on melting curve analysis. All samples were run in triplicate and included a negative control and a positive control. Details of the primers and probes used are provided in **Supplementary Table 2**.

#### *P. malariae* msp-1 genotyping

The *P. malariae msp-1* partial gene amplification was performed on *P. malariae*-positive samples using nested PCR. Amplified products were purified and sequenced to assess genetic diversity. The nested PCR procedure followed a previously established protocol with minor adjustments [28]. In brief, PCR reactions were performed in a 25μL reaction volume, each reaction containing 3μL (∼20ng) of DNA, 300nM of each primer, 2.5 mM MgCl2, 1X PCR buffer, 0.2 mM mixed dNTPs, and 0.5 units of Taq DNA polymerase (Qiagen, Hilden, Germany). This PCR amplicon was used as a template for the nested PCR performed in a 25μL reaction volume, each reaction amplifying *P. malariae msp-1*gene-specific fragments of various sizes. The details of the primers, PCR conditions and amplicon sizes obtained for subsequent sequencing are provided in **Supplementary Table 3**. PCR amplicons were stained with SYBR green while run on a 1.5% gel electrophoresis. The predefined product was visualized with a UV transilluminator. PCR products were purified using Exo-SAP-IT PCR (Applied Biosystems, Beverly, MA, USA) and purified amplicons were used as a sequencing template using the BigDye Terminator v.1.1 Cycle Sequencing Kit on an ABI 3130XL DNA sequencer (Applied Biosystems, Beverly, MA, USA).

### Statistical and phylogenetic analysis

Data were analysed using R software version 4.3.2 (http://www.r-project.org), with variables expressed as proportions for categorical data and medians (with range) for continuous data. The study participants were stratified into three groups: <5, 5-15, and >15 years. Prevalence of *Plasmodium* spp. was calculated and compared across diagnostic methods using appropriate statistical tests. Risk factors associated with infections were examined using logistic regression. Relative risk ratios (RRs) with 95% confidence intervals (CIs) were calculated to determine the risk factors associated with *Plasmodium* infections. Participants with an axillary temperature of 37.5°C or higher were considered to be febrile. A p-value < 0.05 was considered statistically significant.

Sequences were aligned using SeqMan software (https://www.dnastar.com/software), and the resulting sequence contigs were aligned to the published *P. malariae msp-1* (FJ824669) obtained from GenBank database (http://www.ncbi.nlm.nih.gov/GenBank) using the multiple alignment algorithm and MAFFT version 7 using the G-INS-i model. Phylogenetic trees were reconstructed using MEGA version 11 (www.megasoftware.net) [29], employing the Maximum Likelihood method and the General Time Reversible (GTR) plus Gamma Distribution model. The statistical robustness and reliability of the branching order were confirmed via bootstrapping with 1000 replicates. The resulting phylogenetic tree was annotated and visualized using the online tool iTOL v6 (https://itol.embl.de/) [30]. The representative sequences of *P. malariae msp-1* sequenced in this study have been deposited in the NCBI GenBank database and can be retrieved with accession numbers PX662751-PX662813

## Results

### Demographic Profile of the Study Cohort

A cohort of 484 individuals participated in this study, with 392 (81%) enrolled in 2022 and 92 (19%) in 2023. The age range of participants spanned from 3 to 90 years, with a median age of 25 years (ranging from 3 to 90 years) in 2022 and 37 years (ranging from 6 to 72 years) in 2023. The proportion of females exceeded that of males in both years, comprising 62% (n=241) in 2022 and 69% (n=63) in 2023. Among the three defined age groups, individuals older than 15 years constituted the largest segment in both sampling years, comprising 58% (2022; n= 227) and 80% (2023; n= 73) of participants, respectively. A total of 15% (71 out of 484 participants) exhibited fever (≥37.5℃) as their sole malaria symptom at the time of the survey. Additionally, 69% (n=334) were recruited from the Ouidah district, while 31% (n=150) were from the Kpomasse district. All socio-demographic attributes of participants in both 2022 and 2023 are provided in **Table 1**.

**Table 1.** Socio demographic characteristics of the study population.

| Characteristics | DS 2022 (392) | DS (92) | Total (484) |
| --- | --- | --- | --- |
| **Age range (years)** | 3 - 90 | 6 - 72 | 3 - 90 |
| Median (IQR) | 25 (31.3) | 37 (27.3) | 27.5 (33) |
| **BMI (Body mass Index)** | 5.3 - 49.1 | 12.4 - 37.3 | 5.3 - 49.1 |
| Median (IQR) | 18.4 (7.2) | 20.5 (7.6) | 18.7 (7.4) |
| **Gender n (%)** |  |  |  |
| Female | 241 (61.5) | 63 (68.5) | 304 (62.8) |
| Male | 151 (38.5) | 29 (31.5) | 180 (37.2) |
| **Temperature (°C) n (%)** |  |  |  |
| ≤ 37.5°C | 334 (85.2) | 79 (85.9) | 413 (85.3) |
| > 37.5°C | 58 (14.8) | 13 (14.1) | 71 (14.7) |
| **Haemoglobin (g/dl) range** | 6.5 - 18.3 | 6.2 - 16.9 | 6.2 - 18.3 |
| Median (IQR) | 11.9 (1.8) | 11.7 (1.7) | 11.9 (1.7) |
| **Age groups (year) n (%)** |  |  |  |
| < 5 | 14 (3.6) | 1 (1.1) | 15 (3.1) |
| 5 - 15 | 151 (38.5) | 18 (19.6) | 169 (34.9) |
| > 15 | 227 (57.9) | 73 (79.3) | 300 (62) |
| **Location n (%)** |  |  |  |
| Ouidah | 308 (78.6) | 26 (28.3) | 334 (69) |
| Kpomasse | 84 (21.4) | 66 (71.7) | 150 (31) |

DS: Dry Season, n: represents the number of participants and the corresponding percentage value is indicated in brackets, IQR: Interquartile Range

### Prevalence of *Plasmodium* infections by microscopy, RDT, Nested PCR and nested qPCR

The prevalence of *Plasmodium* infections varied according to the diagnostic method used in the 484 samples tested with microscopy, RDT, and nested PCR, and the 302 samples tested with nested qPCR. The overall prevalence of *Plasmodium* infections determined by microscopy was 17% (82/484), while rapid diagnostic tests (RDTs) indicated a slightly higher prevalence of 20% (96/484). However, this difference was not statistically significant (P=0.2). In contrast, nested PCR demonstrated a significantly higher prevalence of 38% (182/484) in comparison to microscopy and RDT (P<0.0001). Furthermore, the utilisation of nested quantitative PCR (qPCR) revealed a 12% increase in *Plasmodium* spp. prevalence in individuals who tested negative using nested PCR.

### Species-specific infection prevalence

Four diagnostic methods were employed to determine the prevalence of *P. falciparum* and non-falciparum infections: microscopy, RDTs, nested PCR, and nested qPCR.

Microscopy detected two *Plasmodium* species (*P. falciparum* and *P. malariae*). Overall, *P. falciparum* infections were identified in 16.7% (81/484) of participants, while *P. malariae* was observed in 0.8% (4/484). Among these infections, 16.1% (78/484) were *P. falciparum* mono-infections and 0.2% (1/484) were *P. malariae* mono-infections (**Table 2**). Furthermore, using RDTs, 19.8% (96/484) of the study population tested positive for *Plasmodium* spp. Among these cases, *P. falciparum* accounted for 19.4% (94/484), while 0.4% (2/484) were classified as *P. ovm* (*P. ovale/P. vivax/P. malariae*) infections (**Table 3**).

**Table 2.** Prevalence of *Plasmodium* infections in Southern Benin population detected by microscopy, Nested PCR and Nested qPCR.

| *Plasmodium* spp. infections | Microscopy (n= 484) |  | Nested PCR (n= 484) |  | Nested qPCR (n=302) |  |
| --- | --- | --- | --- | --- | --- | --- |
|  | N | Prevalence (%) | N | Prevalence (%) | N | Prevalence (%) |
| **All *Plasmodium* Positive** | 82 | 17 | 182 | 37.6 | 36 | 11.9 |
| *Pf* | 81 | 16.7 | 172 | 35.5 | 20 | 6.6 |
| *Pm* | 4 | 0.8 | 32 | 6.6 | 25 | 8.2 |
| *Po* | 0 | 0 | 5 | 1 | 0 | 0 |
| **Mono-infections** | 79 | 16.3 | 157 | 32.4 | 27 | 8.9 |
| *Pf* | 78 | 16.1 | 147 | 30.4 | 11 | 3.6 |
| *Pm* | 1 | 0.2 | 10 | 2.1 | 16 | 5.3 |
| *Po* | 0 | 0 | 0 | 0 | 0 | 0 |
| **Co-infection** | 3 | 0.6 | 25 | 5.2 | 9 | 3 |
| *Pf* + *Pm* | 3 | 0.6 | 20 | 4.1 | 9 | 3 |
| *Pf* + *Po* | 0 | 0 | 3 | 0.6 | 0 | 0 |
| *Pf* + *Pm* + *Po* | 0 | 0 | 2 | 0.4 | 0 | 0 |

*Pf*: *P. falciparum*, *Pm*: *P. malariae*, *Po* : *P. ovale*
N: Number of participants positive for each *Plasmodium* spp., *Pf* infections: mono and mixed *P. falciparum* infections, *Pm* infections: mono and mixed *P. malariae* infections, *Po* infections: mono and mixed *P. malariae* infections

**Table 3.**
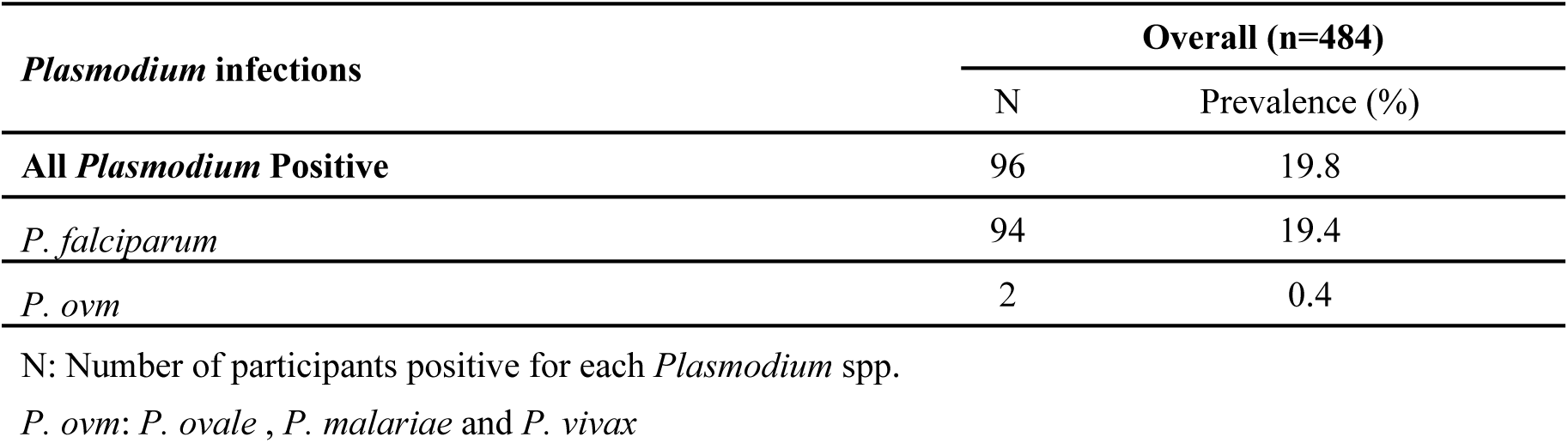
Prevalence of *Plasmodium* spp. in Southern Benin population using RDTs.

Nested PCR identified three *Plasmodium* spp. (*P. falciparum*, *P. malariae* and *P. ovale* spp.). *P. falciparum* and *P. malariae* in 35.5% (172/484) and 6.6% (32/484) of the study population, respectively **(Table 2**). This represents a significant increase compared to microscopy (P < 0.0001). The prevalences observed increased by 6.6% for *P. falciparum* and 8.2% for *P. malariae* when using nested qPCR compared to nested PCR. The *P. ovale* species were observed exclusively as co-infections. It is noteworthy that two participants were found to be triply infected with *P. falciparum*, *P. malariae* and *P. ovale walikeri* (data not shown). Furthermore, no isolates were found to be positive for *P. vivax*.

### ub-microscopic *Plasmodium* infections prevalence and predictors

Sub-microscopic infections were assessed for *Plasmodium* spp. and specifically *P. malariae*, and the factors associated with these types of infections in our sample population.

Sub-microscopic *Plasmodium* infections were detected in 37% of our study population. Specifically, sub-microscopic *P. malariae* infections were detected in 11.2% of participants, of which 6% were mono-infections and 5.2% were mixed infections (**Supplementary Table 4**). The risk of *Plasmodium* infections among Ouidah residents was significantly increased for sub-microscopic infections (RR: 1.5, 95% CI: 1.1-2.2; P = 0.01) (**Table 4**). In addition, *P. malariae* infection indicated a marginal risk of *P. malariae* infection in females compared to males (RR: 1.3, 95% CI: 0.9-8; P = 0.04) (**Table 4**). No significant risk factors were identified for other socio-demographic characteristics.

**Table 4.** Association of socio-demographic characteristics with sub-microscopic *Plasmodium* infections and *P. malariae*.

| Characteristics | Total number | *Plasmodium* spp. |  | P- value | *P. malariae* |  | P- value |
| --- | --- | --- | --- | --- | --- | --- | --- |
| overall | 402 | n (%) | RR (95% CI) |  | n (%) | RR (95% CI) |  |
| **Gender** |  |  |  |  |  |  |  |
| Female | 258 | 95 (36.8) | 1 (0.8 - 1.3) | 0.9 | 20 (7.8) | 1.3 (0.9 -- 8) | 0.04 |
| Male | 144 | 53 (36.8) |  |  | 4 (2.8) |  |  |
| **Age groups** |  |  |  |  |  |  |  |
| < 5 | 15 | 7 (46.7) | Reference |  | 1 (6.7) | Reference |  |
| 5 -- 15 | 112 | 47 (42) | 0.9 (0.5 - 1.6) | 0.7 | 4 (4.5) | 0.5 (0.06 - 4.4) | 0.5 |
| > 15 | 275 | 94 (34.2) | 0.7 (0.4 - 1.2) | 0.3 | 19 (7.3) | 1 (0.1 - 7.2) | 0.9 |
| **Temperature (°C)** |  |  |  |  |  |  |  |
| ≤ 37.5°C | 342 | 127 (37.1) | 0.9 (0.6 - 1.3) | 0.7 | 21 (6.1) | 0.8 (0.2 - 2.6) | 0.7 |
| > 37.5°C | 60 | 21 (35) |  |  | 3 (5) |  |  |
| **Location** |  |  |  |  |  |  |  |
| Ouidah | 300 | 121 (40.3) | 1.5 (1.1 - 2.2) | 0.01 | 18 (6) | 1 (0.4 - 2.4) | 0.9 |
| Kpomasse | 102 | 27 (26.5) |  |  | 6 (5.9) |  |  |

n : represents the number of participants positive for each *Plasmodium* spp. and the corresponding prevalence is indicated in brackets

### *P. malariae* msp-1 fragments genetic diversity

The genetic diversity of *P. malariae* was evaluated using a set of 57 samples that were positive for either *P. malariae* mono-infections or co-infections. This diversity was investigated by splitting the *msp-1* gene into five fragments to cover more than half of the gene.

In our samples, fragment 1 encodes a peptide of 177 residues, spanning amino acids 180 to 356 in the unaligned MM1 sequence (GenBank #FJ824669). Notably, polymorphism was observed exclusively within the region of amino acids 231 to 262 (**Fig 1**), with thirty different amino acid sequences identified within this fragment for all samples sequenced. Due to the presence of a microsatellite and numerous non-specific bands after amplification, fragment 2 could not be sequenced. Fragment 3 corresponds to amino acids 970 to 1179 (210 aa) in the MM1 sequence. An insertion of 9 amino acids (PQPQAALPA) relative to the MM1 isolate was detected in ten of our sequences at the position 997 (**Fig 1**). For fragment 4, aligned to amino acids 1380 to 1526 (147 aa) in the MM1 sequence, a deletion of PIVG amino acids at positions 1381 to 1384 and two other substitutions (E1416D in two samples and K1510I in one sample) were observed (**Fig 1**).

**Fig 1.**
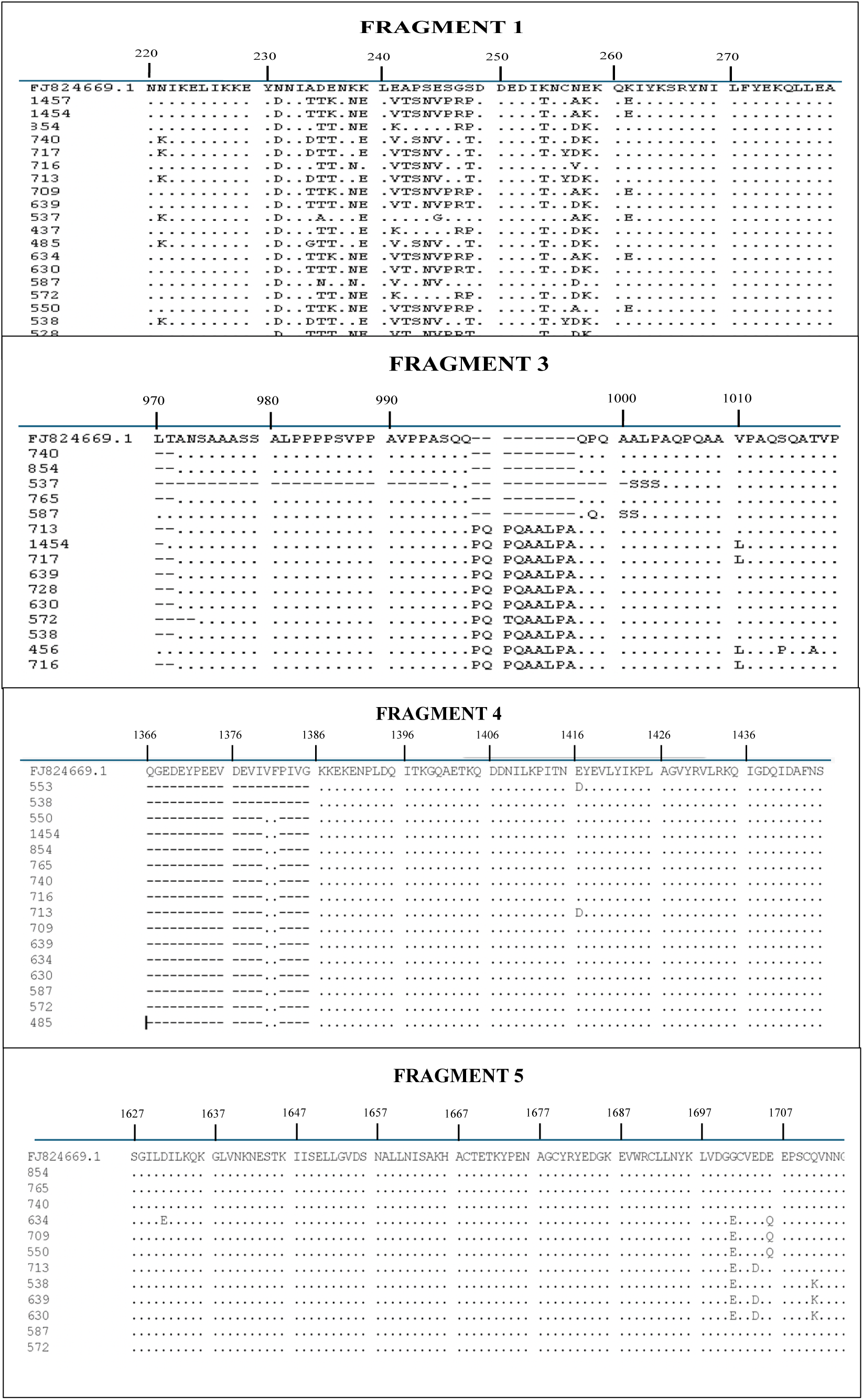
Amino acid polymorphism in the *P. malariae* merozoite surface protein 1 sequences of fragments 1, 3, 4 and 5

In the C-terminal region, fragment 5 was amplified, corresponding to amino acids 1548 to 1747 (200 aa) in the MM1 sequence. Its variability was limited to 8 amino acid substitutions (**Fig 1**). Phylogenetic analysis for msp-1 revealed that *P. malariae* strains from Benin were closely related to the reference strain, indicating their similarity to this reference strain. (**Fig 2).** The results of the nucleotide diversity metrics, including θ (0.008053) and π (0.006015), provide quantitative measures of genetic diversity and indicate variation within the *P. malariae* parasite in the study population (**data not shown**).

**Fig 2.**
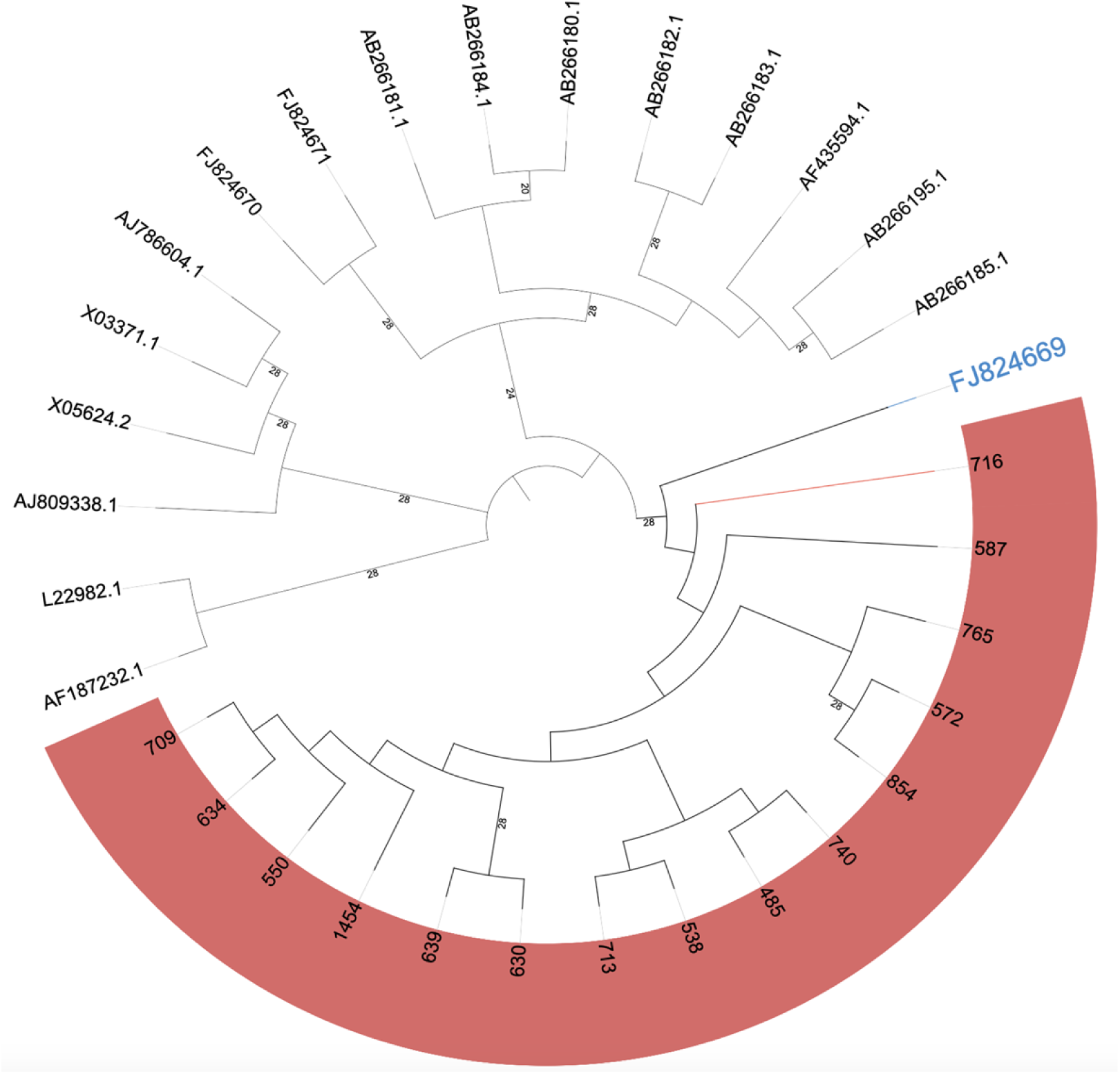
Phylogenetic tree of the MSP-1 gene using the GTR+G+I model, with 17 sequences from 16 different *Plasmodium* species. The *P. malariae MSP-1* sequences from our isolates (in pink) and the *P. malariae MSP-1* reference in blue, and the remaining are references from the other *Plasmodium* species used.

## Discussion

The dominance of *P. falciparum* within *Plasmodium* species in sub-Saharan Africa has led to its prioritization in malaria research, often overshadowing the study of non-falciparum species, particularly in endemic areas like Benin. Comprehensive spatial and temporal surveillance of all *Plasmodium* species, particularly *P. malariae*, among asymptomatic populations, alongside an understanding of its genetic diversity, is crucial for unravelling various facets of malaria research, including epidemiology, transmission dynamics, and evolutionary processes. This study aimed to monitor asymptomatic *Plasmodium* infections and characterize the polymorphisms of the msp-1of *P. malariae* in Beninese individuals. These data remain essential for targeted strategies for effectively controlling and ultimately eradicating malaria.

Community surveillance in Ouidah and Kpomasse, southern Benin, revealed a *Plasmodium* infections rate of 45% in the study area, using molecular methods. This prevalence, while high, is consistent with previous studies from the region[14,24,31], albeit lower than infection rates reported in other West African countries such as Nigeria and Ghana [32,33]. Historically, southern Benin has been classified as a high malaria transmission area, with several contributing factors, including a high entomological inoculation rate (EIR) and suboptimal coverage of prevention measures. The large proportion of asymptomatic carriers in these communities poses a major challenge to malaria control efforts, as these individuals serve as reservoirs for ongoing transmission [34–36]. This underscores the need for community-based strategies that specifically target asymptomatic carriers across all *Plasmodium* species, which are essential for malaria elimination [37,38].

The results of our study confirm that *P. falciparum* is the predominant *Plasmodium* species in the study area [24,39,40]. Notably, *P. malariae* emerged as the second most identified species by microscopy, although its prevalence is less than 1%. Rapid diagnostic tests (RDTs) revealed that only two samples tested positive for *P. ovm*. In contrast, the utilisation of more sensitive diagnostic methodologies, such as nested polymerase chain reaction (nested PCR) and nested quantitative polymerase chain reaction (nested qPCR), has led to a notable increase in the detection of *P. malariae*, with rates of 7% and 12%, respectively. This indicates that conventional microscopy significantly underestimates *P. malariae* infections by up to 11%. A similar rate of sub-microscopic *P. malariae* infection has been reported in a previous study [41].

These findings highlight the challenges encountered in accurately detecting certain parasites using conventional diagnostic tools. Both microscopy and RDTs although widely used in routine malaria diagnosis, have limited sensitivity in detecting *P. malariae*. This is partly due to the parasite’s ability to maintain low parasitaemia in its host, rendering it susceptible to misdiagnosis, particularly when co-infecting with *P. falciparum* [8,42]. The persistence of sub-microscopic *P. malariae* infections can result in the development of latent health complications and increased mortality, emphasising the critical importance of accurate diagnosis. The findings of this study highlight the necessity of incorporating more sensitive diagnostic modalities, such as molecular techniques, to more accurately detect and understand the prevalence of *Plasmodium* parasites beyond *P. falciparum*. It is therefore essential to improve diagnostic accuracy to guide effective treatment strategies and to mitigate the long-term health consequences of undetected malaria infections.

The genetic diversity of *P. malariae* was evaluated using a set of 57 samples that were positive for *P. malariae* mono-infections or co-infections. The *msp-1* gene fragments employed for the investigation of genetic diversity were constructed in a manner that encompassed over 50% of the gene sequence. While five fragments of this gene were targeted for amplification, sequencing of fragment 2 was hindered due to the presence of excessive non-specific bands. Additionally, the amplification of the four other fragments used to study the genetic diversity of this gene was unsuccessful in some samples. This failure may be attributed to low parasitaemia levels of *P. malariae*, as most samples infected with *P. malariae* were detected using highly sensitive molecular techniques. An analysis of the merozoite surface protein 1 (msp-1), a critical vaccine target, revealed nucleotide sequence similarities ranging from 95% to 100% across the four sequenced fragments. This high level of similarity was observed not only with strains from Africa but also with those from another continent (South America), indicating a low genetic diversity of this parasite across the world. These findings are consistent with previously published data indicating a low-level genetic diversity of *P. malariae* [17,28,43,44]. The low genetic diversity of *P. malariae* in the study region may reflect genetic stability across other regions of Benin, although further verification is required, using larger sample sizes and diverse geographical regions. It is noteworthy that a similarly low level of sequence diversity was observed in other non-falciparum species, including *P. ovale* spp. [45]. Conversely, significant genetic diversity in *P. falciparum* has been reported across Africa, including in our study region [46]. The presence of distinct amino acid sequences observed emphasises the significance of genetic diversity in elucidating the biology, transmission dynamics, and disease outcomes of the parasite. The results of the nucleotide diversity metrics, including θ (0.008053) and π (0.006015), provide quantitative measures of genetic diversity and indicate variation within the *P. malariae* parasite in the study population. Future studies should explore the genetic variation observed and its implications for drug resistance, immune evasion, and pathogenicity in *P. malariae* infections.

The results of this study highlight the necessity to expand research efforts beyond *P. falciparum* to address the challenges posed by non-falciparum species such as *P. malariae*. Enhancing rapid diagnostic tests for non-falciparum species, improving microscopist training, and integrating molecular techniques into routine diagnosis are essential steps. Additionally, a better understanding of malaria parasite diversity, transmission dynamics, and evolutionary biology will facilitate the development of more effective strategies for malaria control and eradication. Furthermore, the low genetic diversity and highly conserved amino acid sequences in *msp-1* suggest it may be a promising vaccine candidate for *P. malariae*.

## Supporting information

**S1 Table.** Names and sequences of primers for *Plasmodium* spp. characterization by nested PCR

**S2 Table.** Names and sequences of primers and probes for *Plasmodium* spp. characterization by nested qPCR

**S3 Table.** Names and sequences of primers for *P. malariae msp-*1 amplification by nested PCR

**S4 Table.** Comparison of the microscopic and sub-microscopic prevalence of *Plasmodium* spp.

## Data Availability

All data generated or analysed during this study are included in the article. A total of 63 high-quality sequences corresponding to the amplified fragments of *P. malariae msp-1* gene were submitted to the NCBI GenBank database under accession numbers PX662751-PX662813

## Acknowledgements

We are grateful to the study communities for their participation, and to the Ouidah and Kpomasse health district administrations for their support. We thank Le Thi Kieu Linh for supporting with sequencing procedures and Pulcherie Assogba for microscopy procedures. We also acknowledge the support of the COMAL consortium for funding the sample collection used in this study.

## Author contributions

RA conceptualized the study design and contributed to the study materials for experimental investigations, performed experimental procedures, statistical and phylogenetic analysis, wrote the first draft and acquired the funding. TPV, AAA supervised RA, associated study objectives. SR, VMN, MMN supported RA and performed partial experimental procedures. RAk was involved in the sampling, and microscopy procedures. RAk, SR, VMN, MMN, AAA and TPV reviewed the circulated draft. All authors have read and approved the manuscript.

